# Role of antibodies, inflammatory markers, and echocardiographic findings in post-acute cardiopulmonary symptoms after SARS-CoV-2 infection

**DOI:** 10.1101/2021.11.24.21266834

**Authors:** Matthew S. Durstenfeld, Michael J. Peluso, J. Daniel Kelly, Sithu Win, Shreya Swaminathan, Danny Li, Victor M. Arechiga, Victor Zepeda, Kaiwen Sun, Shirley Shao, Christopher Hill, Mireya I. Arreguin, Scott Lu, Rebecca Hoh, Viva Tai, Ahmed Chenna, Brandon C. Yee, John W. Winslow, Christos J. Petropoulos, John Kornak, Timothy J. Henrich, Jeffrey N. Martin, Steven G. Deeks, Priscilla Y. Hsue

## Abstract

**BACKGROUND:** Shortness of breath, chest pain, and palpitations occur as post-acute sequelae of COVID-19 (PASC), but whether symptoms are associated with echocardiographic abnormalities, cardiac biomarkers, or markers of systemic inflammation remains unknown.

**METHODS:** In a cross-sectional analysis, we assessed symptoms, performed echocardiograms, and measured biomarkers among adults >8 weeks after PCR-confirmed SARS-CoV-2 infection. We modeled associations between symptoms and baseline characteristics, echocardiographic findings, and biomarkers using logistic regression.

**RESULTS:** We enrolled 102 participants at a median 7.2 months (IQR 4.1-9.1) following COVID-19 onset; 47 individuals reported dyspnea, chest pain, or palpitations. Median age was 52 years (range 24-86) and 41% were women. Female sex (OR 2.55, 95%CI 1.13-5.74) and hospitalization during acute infection (OR 3.25, 95%CI 1.08-9.82) were associated with symptoms. IgG antibody to SARS-CoV-2 receptor binding domain (OR 1.38 per doubling, 95%CI 1.38-1.84) and high-sensitivity C-reactive protein (OR 1.31 per doubling, 95%CI 1.00-1.71) were associated with symptoms. Regarding echocardiographic findings, 4/47 (9%) with symptoms had pericardial effusions compared to 0/55 without symptoms (p=0.038); those with pericardial effusions had a median 4 symptoms compared to 1 without (p<0.001). There was no strong evidence for a relationship between symptoms and echocardiographic functional parameters (including left ventricular ejection fraction and strain, right ventricular strain, pulmonary artery pressure) or high-sensitivity troponin, NT-pro-BNP, interleukin-10, interferon-gamma, or tumor necrosis factor-alpha.

**CONCLUSIONS:** Among adults in the post-acute phase of SARS-CoV-2 infection, SARS-CoV-2 RBD antibodies, markers of inflammation and, possibly, pericardial effusions are associated with cardiopulmonary symptoms. Investigation into inflammation as a mechanism underlying PASC is warranted.

**FUNDING:** This work was supported by the UCSF Division of Cardiology at Zuckerberg San Francisco General, and the National Institutes of Health/National Heart Lung Blood Institute and National Institute of Allergy and Infectious Diseases. MSD is supported by NIH 5K12HL143961. MJP is supported on NIH T32 AI60530-12. JDK is supported by NIH K23AI135037. TJH is supported by NIH/NIAID 3R01A1141003-03S1. PYH is supported by NIH/NAID 2K24AI112393-06. This publication was supported by the National Center for Advancing Translational Sciences, National Institutes of Health, through UCSF-CTSI Grant Number UL1TR001872. Its contents are solely the responsibility of the authors and do not necessarily represent the official views of the NIH.

**GRAPHICAL ABSTRACT:** 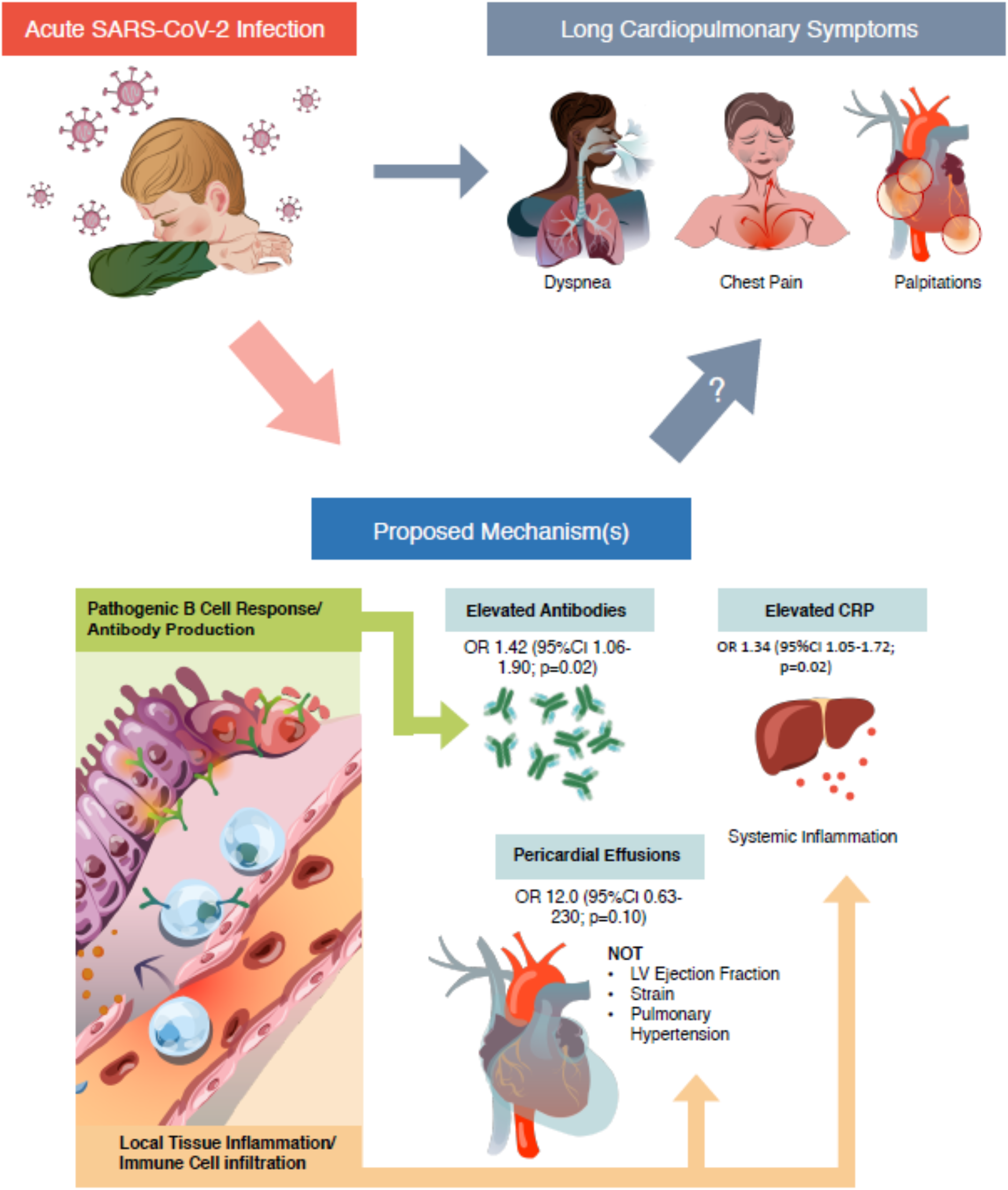

## Introduction

Following acute SARS-CoV-2 infection, a significant proportion of individuals have shortness of breath, chest pain, and palpitations (hereafter referred to as cardiopulmonary symptoms, as shorthand) that persist for at least 12 months (1-3). Studies performed during early convalescence suggest that cardiac inflammation and fibrosis may be evident by cardiac magnetic resonance (CMR) imaging in some individuals 2-3 months after acute COVID-19 (4-6). In contrast, short-term transthoracic echocardiography (TTE) studies of patients with COVID-19 after hospital discharge have not found cardiac abnormalities compared to matched controls, even among those with biochemical evidence of myocardial injury while hospitalized (7). Thus, whether persistent cardiopulmonary symptoms in “Long COVID” or post-acute sequelae of COVID-19 (PASC) are explained by cardiac structural or functional changes or part of a systemic inflammatory post-viral syndrome has become a major clinical question.

If cardiac pathology underlies PASC, TTE may provide clues regarding mechanisms of ongoing symptoms. To date, studies published have not investigated the link between persistent cardiopulmonary symptoms attributed to “Long COVID” and cardiac structural or functional changes beyond the early recovery phase (e.g., > 2 months). Another major limitation of the existing literature is that most studies have included only those with severe COVID-19 requiring hospitalization, while most individuals with COVID-19 are not hospitalized. One study that examined cardiac changes 6 months after mild infection among healthcare workers using CMR had few participants with persistent symptoms (8). In April 2020, a prospective cohort of individuals with documented SARS-CoV-2 infection was established in Northern California (Long-term Impact of Infection with Novel Coronavirus, LIINC, www.liincstudy.org), which includes individuals with asymptomatic to severe disease. Therefore, the objective of this study was to determine whether echocardiographic findings, cardiac biomarkers, and inflammatory biomarkers obtained months after acute COVID-19 are associated with persistent cardiopulmonary symptoms.

## Methods

### Design

Cross-sectional evaluation of a prospective cohort study.

### Participants

As previously described, the LIINC COVID-19 recovery cohort (NCT04362150) was established in April 2020 to study the effects of infection with SARS-CoV-2 (9). Most participants were recruited from the general community with some referred from acute studies. Confirmed SARS-CoV-2 infection with documentation of nucleic acid amplification testing was required. Individuals were queried regarding the presence of 32 individual symptoms from the Centers for Disease Control list of COVID-19 symptoms and from the Patient Health Questionnaire Somatic Symptom Scale (10). A symptom was considered present if it was new in onset or worsened (if pre-existing) since the time of SARS-CoV-2 infection. We invited the first 95 participants who sequentially indicated interest in participating without respect to symptom status and enrolled all eligible and willing individuals (n=78); thereafter, we continued to enroll all eligible people living with HIV (n=13) and selectively enrolled those without HIV who reported symptoms at their prior LIINC study visit (n=11). We excluded pregnant individuals and those with history of heart failure, pulmonary hypertension, moderate or severe valvular disease, congenital heart disease, or organ transplant prior to COVID-19.

### Measurements

#### Questionnaire-based

Participants completed a structured interview about medical history prior to SARS-CoV-2 infection, characteristics of acute infection, cardiopulmonary diagnoses, and symptoms within the previous two weeks, which were considered potentially COVID-19-related only if new or worsened. We systematically asked about fatigue, shortness of breath, chest pain, palpitations, syncope, and edema. After reports of postural orthostatic tachycardia syndrome (POTS) we added questions about positional symptoms (11).

#### Echocardiographic

We measured supine blood pressure. A blinded cardiac sonographer performed echocardiograms using a standardized protocol with a GE VIVID E90 machine. Echocardiograms were measured and post-processed by a single echocardiographer with GE EchoPAC software. Volumes and ejection fraction were measured using the modified Simpson’s rule using the biplane method of discs and indexed to body surface area.(12) Diastolic function was assessed according to the 2016 American Society of Echocardiography (ASE) guidelines (13). Additional quantitative parameters were measured including early to late diastolic filling ratio (E/A), early diastolic lateral and medial mitral annulus tissue doppler velocity (e’), early diastolic filling to tissue velocity ratio (E/e’), tricuspid annular plane systolic excursion (TAPSE), and right ventricular systolic excursion velocity (S’). Pulmonary artery pressures were estimated based on tricuspid regurgitant continuous wave Doppler tracings using the modified Bernoulli equation and right atrial pressure estimated by inferior vena cava size and collapsibility. Cardiac index was estimated using left ventricular outflow tract velocity time integrals and diameter indexed to body surface area. Left ventricular peak systolic global longitudinal strain, peak systolic dispersion, and right ventricular free wall peak systolic strain were measured using focused ventricular views at greater than 60 frames per second. Myocardial work, constructive work, wasted work, and work efficiency were measured based on strain measurements and non-invasive blood pressure. Presence of pericardial effusion was assessed from multiple views and confirmed by a second, blinded echocardiographer.

#### Blood-based

Venous blood was collected on the day of the echocardiogram and processed for serum and plasma. Samples were batch processed for measurement of high sensitivity troponin I (hs-trop; ADVIA Centaur® High-Sensitivity Troponin I (TNIH) assay), high sensitivity c-reactive protein (hs-CRP; ADVIA® Chemistry CardioPhase™ High Sensitivity C-Reactive Protein assay), and N-terminal prohormone b-type natriuretic protein (NT-pro-BNP; Roche Cobas 6000 Elecsys® proBNP II assay). Additional markers including interleukin 6 (IL-6), interleukin 10 (IL-10), interferon gamma (IFNγ), tumor necrosis factor alpha (TNFα), and SARS-CoV-2 receptor binding domain IgG according were measured on a subset of participants (n=73) with specimens collected 90-160 days after symptom onset and analyzed by Monogram Biosciences using the Quanterix Simoa® platform. Samples were assayed blinded with respect to patient and clinical information. Assay performance was consistent with the manufacturer’s specifications.

### Statistical Analysis

As our primary outcome of interest was the presence of cardiopulmonary PASC, our primary analyses consisted of logistic regression models first for the composite outcome of presence of dyspnea, chest pain, or palpitations, then for presence of each individual symptom. As a proxy for more severe PASC, we assessed the distribution of the number of symptoms reported to compare the quartile with the most symptoms with the quartile with the fewest symptoms; therefore, we modeled having two or more symptoms relative to no symptoms (14). We modeled associations between presence of cardiopulmonary symptoms (outcome) and demographic/clinical variables including race/ethnicity, educational attainment, income, body mass index, past medical history, and acute COVID-19 severity markers (exposures) and included age and sex as confounders. We used the same approach to model associations between symptoms and echocardiographic parameters, including age, sex, hospitalization, and time since symptom onset as confounders. We included past medical history in adjusted models where pertinent, specifically including hypertension for LV mass index, left atrial volume, and diastolic dysfunction, and lung disease for right ventricular parameters. Similarly, we included HIV as a covariate when supported by prior literature and assessed for interaction between HIV and LV mass index (15), diastolic function (15, 16), LV strain (17, 18), and pulmonary artery pressures (19). We followed the same approach to model the association between symptoms and biomarkers, which were natural log-transformed due to right-skewed distributions. We assessed for interactions between sex and troponin and NT-pro-BNP and interactions between HIV and inflammatory markers (hsCRP, IL-6, etc). To model associations between symptoms and echocardiographic parameters where there was complete separation of the variable of interest in the fitted logistic regression model (i.e., pericardial effusions), we used Firth logistic regression (20). To model correlation between antibodies and inflammatory markers, we used linear regression of log-transformed biomarkers. Data were recorded using REDCap. Statistical analyses were performed using StataMP 16.1 (StataCorp, College Station, TX).

### Study Approval

All participants provided signed written informed consent prior to participation. Institutional Review Board approval was granted by the University of California, San Francisco. Dr. Durstenfeld had full access to the study data and takes responsibility for the data integrity and analysis.

## Results

### Clinical Characteristics among Individuals with and without Persistent Cardiopulmonary Symptoms following COVID-19

Of the 115 people we contacted, six did not respond, four declined to participate, two screened out (pregnancy and congenital heart disease) and one participant dropped out after signing the consent but before completing a study visit; therefore, 102 participants completed a study visit with an echocardiogram from November 2020-May 2021 at a median of 7.2 months (IQR 4.1-9.1) after SARS-CoV-2 infection defined as symptom onset or positive PCR testing among those with asymptomatic infection. As shown in Table 1, those with dyspnea, chest pain, or palpitations differed from those without those symptoms with respect to sex, body mass index, and hospitalization for acute infection.

**Table 1.**
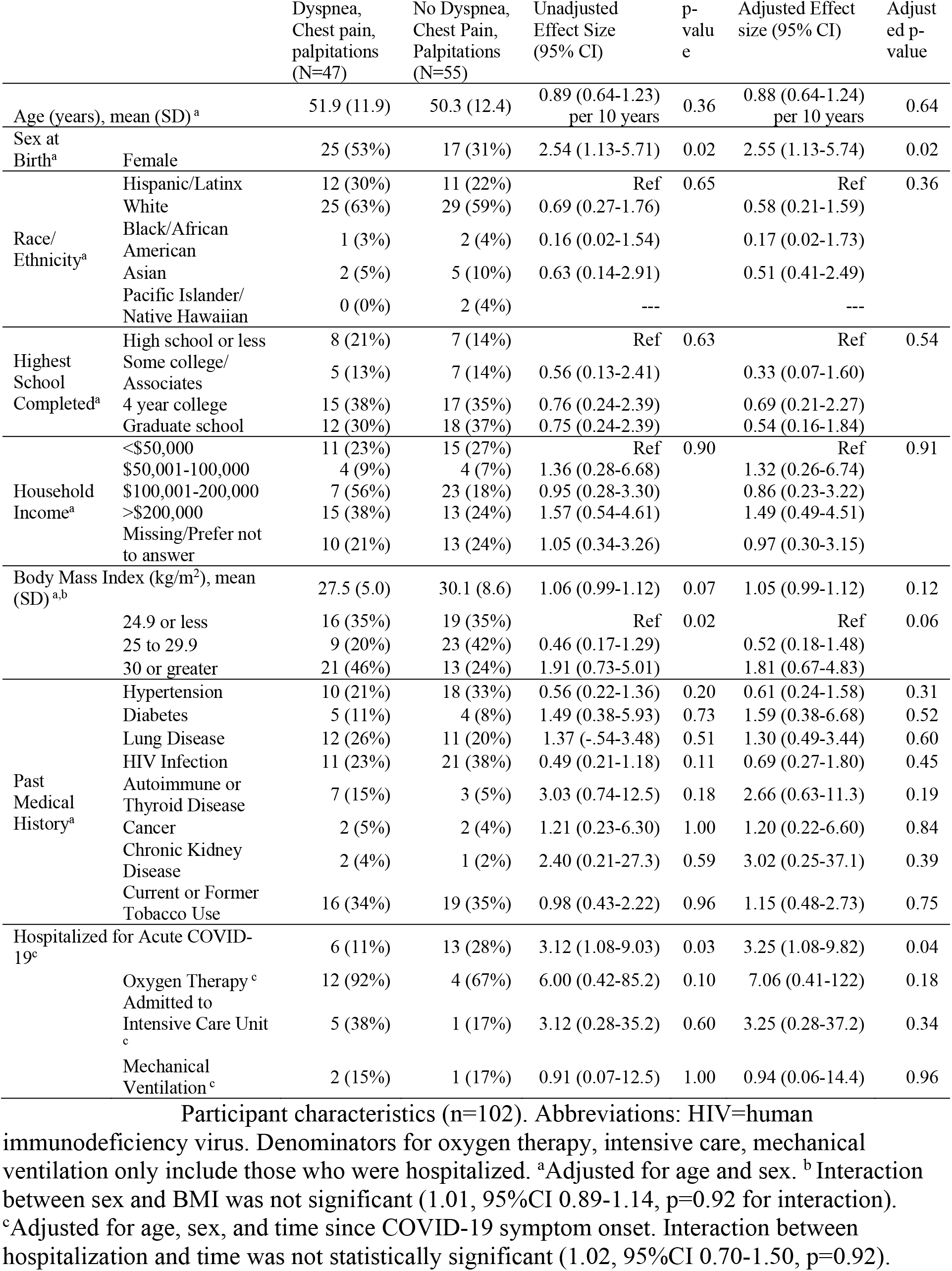
Demographics and Past Medical History Stratified by Symptoms (N=102)

Of the 102 participants, 64 had at least one potentially cardiopulmonary symptom including dyspnea, chest pain, palpitations, fatigue, edema, syncope, or postural symptoms. Of those, 47 individuals had the composite primary outcome of dyspnea, chest pain or palpitations in the preceding two weeks; 33 had dyspnea, 15 had chest pain, and 27 had palpitations. Fatigue and dyspnea were most common (Figure 1), and 50% reported a reduction in exercise capacity. Nearly all with dyspnea characterized their dyspnea as exertional (31/33, 94%), but few participants with chest pain reported that their chest pain was associated with activity (3/15, 20%). Palpitations were universally paroxysmal, and some participants reported sustained tachycardia after exercise or with position change. The median number of symptoms was 1 (IQR 0-2, range 0-6); 38 individuals reported no symptoms and 38 reported 2 or more symptoms. Symptoms remained persistent over time following acute infection (Supplemental Figure 1).

**Figure 1.**
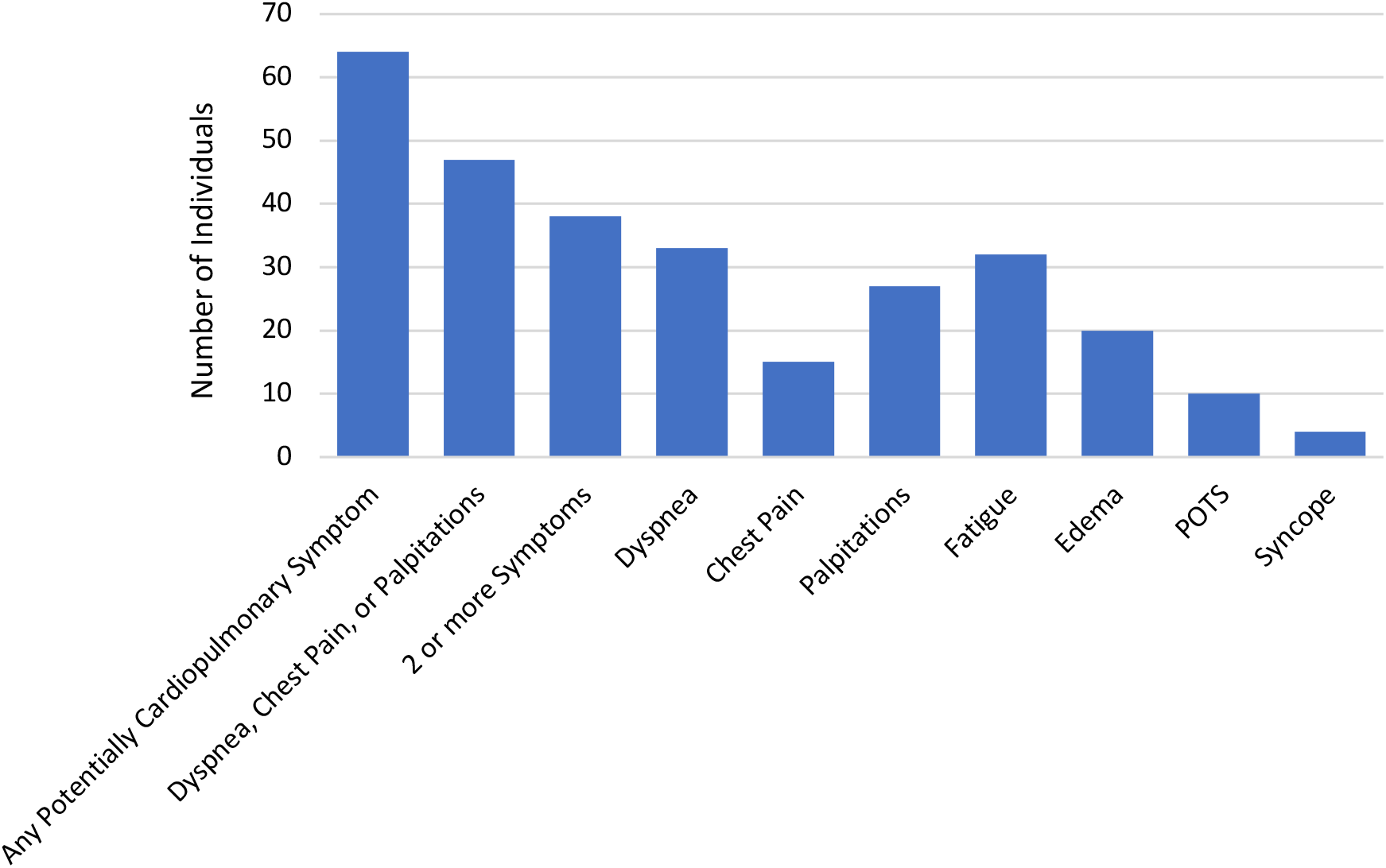
Cardiopulmonary Symptoms Represented in Study Sample (N=102) Number of participants who reported potentially cardiopulmonary symptoms in the 2 weeks prior to echocardiogram at a median of 7.2 months (IQR 4.1-9.1) after COVID-19 onset (N=102). These are not prevalence estimates within the population of those recovering from COVID-19, but are presented to provide context for the associations between echocardiographic findings, biomarkers, and symptoms. POTS=postural orthostatic tachycardia syndrome, although half of those who reported postural symptoms consistent with POTS had not been formally diagnosed.

### Sociodemographic Characteristics and Severity of Acute Illness Associated with Persistent Symptoms

Female sex (OR 2.54, 95%CI 1.13-5.74; p=0.02) was generally associated with the composite outcome and individual symptoms including dyspnea (OR 2.71, 95%CI 1.16-6.37; p=0.02), chest pain (OR 2.45, 95%CI 0.80-7.52; p=0.12), and palpitations (OR 4.40, 95%CI 1.70-11.4; p=0.002). Hospitalization was similarly associated with the composite outcome (OR 3.25 95% 1.08-9.82; p=0.04) and with dyspnea (OR 3.06, 95%CI 1.04-8.78; p=0.04), chest pain (OR1.80, 95%CI 0.48-6.76; p=0.38), and palpitations (OR 5.45, 95%CI 1.66-18.0; p=0.005). Hypertension and HIV were associated with lower odds of symptoms and chronic kidney disease and autoimmune disease with higher odds as shown in Table 1; the small number of individuals with specific comorbidities precludes drawing strong conclusions regarding the associations between past medical history and symptoms. Similarly, there was a suggestion that oxygen therapy and admission to the intensive care unit may be associated with symptoms, but the sample of hospitalized individuals was too small for a precise estimate of these effects.

### Echocardiographic Findings

The prevalence of reduced cardiac function (LVEF <50%, diastolic dysfunction, LV strain > -18%, RV strain > -20%, or qualitative RV dysfunction) was 36%, and most abnormalities were clinically minor (i.e., mild diastolic dysfunction or mildly reduced strain). Only two individuals had LVEF less than 50%. One individual had been previously diagnosed with a dilated heart (but not clinical heart failure) attributed to HIV and was taking medical therapy. Another individual had a myocardial infarction after infection with SARS-CoV-2 with a regional wall motion abnormality in a coronary artery distribution. Only mild diastolic dysfunction, mild RV dysfunction, mild valvular disease by ACC/AHA guidelines(21) was present, and no one had echocardiographic evidence of pulmonary hypertension.

Among those with the composite outcome, 4/47 (9%) had pericardial effusions compared with 0/55 without symptoms (p=0.038). Pericardial effusions were all trace or small, and none had echocardiographic signs of hemodynamic significance. All individuals with pericardial effusions had normal LVEF, normal strain, normal RV size and function, and normal pulmonary artery pressures; two had mild diastolic dysfunction. One of the four individuals was diagnosed clinically with post-COVID myopericarditis and treated with a non-steroidal anti-inflammatory (diclofenac) and aspirin without improvement; the other three had not been treated with anti-inflammatory medications. Pericardial effusions were highly associated with symptoms with a large estimated odds ratio, but which did not reach statistical significance (OR 12.0, 95%CI 0.63-230; p=0.098). In sensitivity analysis adjusting for autoimmune disease and HIV, the effect estimate was still large, but again did not reach statistical significance (OR 9.2, 95%CI 0.45-187; p=0.15). The median number of symptoms among those with a pericardial effusion was 4 compared to 1 among those without pericardial effusion (p=0.0007; Supplemental Table), and pericardial effusions were associated with estimated odds of having 2 or more symptoms that were 10.7 times higher that did not reach statistical significance (95%CI 0.55-206; p=0.12).

As shown in Table 2, other echocardiographic parameters such as LVEF and LV mass index (Figure 2) were not clearly associated with symptoms with two possible exceptions. Since no participants had more than mild diastolic dysfunction, our data are inconclusive regarding potential association between diastolic dysfunction and the composite outcome (OR 1.77, 95% 0.35-8.88; p=0.78). Secondly, while our data suggest that RV dilation may be associated with increased odds of symptoms, the confidence interval is very wide implying inconclusive results (OR3.55, 95%CI 0.28-45.3; p=0.31). Findings were similar when considering individual symptoms and the presence of 2 or more symptoms compared to no symptoms (Table 3).

**Table 2.**
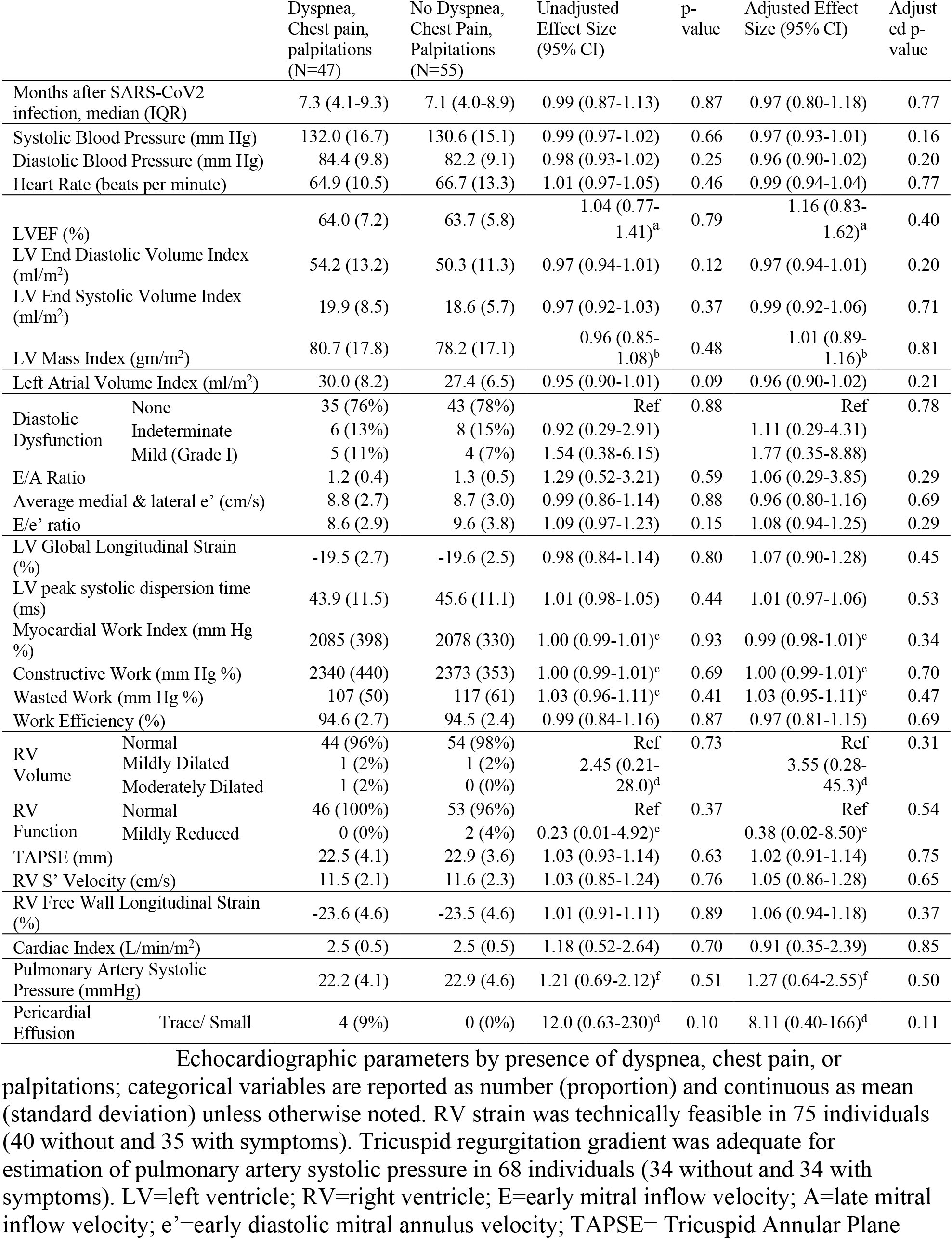
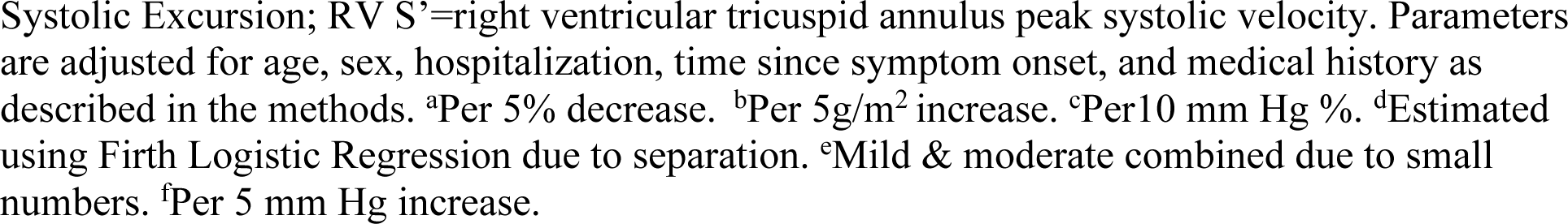
Echocardiographic Parameters and Association with Presence of Dyspnea, Chest Pain or Palpitations.

|  |  | Dyspnea,<br>Chest pain,<br>palpitations<br>(N=47) | No Dyspnea,<br>Chest Pain,<br>Palpitations<br>(N=55) | Unadjusted<br>Effect Size<br>(95% CI) | p-<br>value | Adjusted Effect<br>Size (95% CI) | Adjust<br>ed p-<br>value |
| --- | --- | --- | --- | --- | --- | --- | --- |
| Months after SARS-CoV2<br>infection, median (IQR) |  | 7.3 (4.1-9.3) | 7.1 (4.0-8.9) | 0.99 (0.87-1.13) | 0.87 | 0.97 (0.80-1.18) | 0.77 |
| Systolic Blood Pressure (mm Hg) |  | 132.0 (16.7) | 130.6 (15.1) | 0.99 (0.97-1.02) | 0.66 | 0.97 (0.93-1.01) | 0.16 |
| Diastolic Blood Pressure (mm Hg) |  | 84.4 (9.8) | 82.2 (9.1) | 0.98 (0.93-1.02) | 0.25 | 0.96 (0.90-1.02) | 0.20 |
| Heart Rate (beats per minute) |  | 64.9 (10.5) | 66.7 (13.3) | 1.01 (0.97-1.05) | 0.46 | 0.99 (0.94-1.04) | 0.77 |
| LVEF (%) |  | 64.0 (7.2) | 63.7 (5.8) | 1.04 (0.77-1.41)<sup>a</sup> | 0.79 | 1.16 (0.83-1.62)<sup>a</sup> | 0.40 |
| LV End Diastolic Volume Index<br>(ml/m<sup>2</sup>) |  | 54.2 (13.2) | 50.3 (11.3) | 0.97 (0.94-1.01) | 0.12 | 0.97 (0.94-1.01) | 0.20 |
| LV End Systolic Volume Index<br>(ml/m<sup>2</sup>) |  | 19.9 (8.5) | 18.6 (5.7) | 0.97 (0.92-1.03) | 0.37 | 0.99 (0.92-1.06) | 0.71 |
| LV Mass Index (gm/m<sup>2</sup>) |  | 80.7 (17.8) | 78.2 (17.1) | 0.96 (0.85-1.08)<sup>b</sup> | 0.48 | 1.01 (0.89-1.16)<sup>b</sup> | 0.81 |
| Left Atrial Volume Index (ml/m<sup>2</sup>) |  | 30.0 (8.2) | 27.4 (6.5) | 0.95 (0.90-1.01) | 0.09 | 0.96 (0.90-1.02) | 0.21 |
| Diastolic<br>Dysfunction | None | 35 (76%) | 43 (78%) | Ref | 0.88 | Ref | 0.78 |
|  | Indeterminate | 6 (13%) | 8 (15%) | 0.92 (0.29-2.91) |  | 1.11 (0.29-4.31) |  |
|  | Mild (Grade I) | 5 (11%) | 4 (7%) | 1.54 (0.38-6.15) |  | 1.77 (0.35-8.88) |  |
| E/A Ratio |  | 1.2 (0.4) | 1.3 (0.5) | 1.29 (0.52-3.21) | 0.59 | 1.06 (0.29-3.85) | 0.29 |
| Average medial & lateral e' (cm/s) |  | 8.8 (2.7) | 8.7 (3.0) | 0.99 (0.86-1.14) | 0.88 | 0.96 (0.80-1.16) | 0.69 |
| E/e' ratio |  | 8.6 (2.9) | 9.6 (3.8) | 1.09 (0.97-1.23) | 0.15 | 1.08 (0.94-1.25) | 0.29 |
| LV Global Longitudinal Strain<br>(%) |  | -19.5 (2.7) | -19.6 (2.5) | 0.98 (0.84-1.14) | 0.80 | 1.07 (0.90-1.28) | 0.45 |
| LV peak systolic dispersion time<br>(ms) |  | 43.9 (11.5) | 45.6 (11.1) | 1.01 (0.98-1.05) | 0.44 | 1.01 (0.97-1.06) | 0.53 |
| Myocardial Work Index (mm Hg<br>%) |  | 2085 (398) | 2078 (330) | 1.00 (0.99-1.01)<sup>c</sup> | 0.93 | 0.99 (0.98-1.01)<sup>c</sup> | 0.34 |
| Constructive Work (mm Hg %) |  | 2340 (440) | 2373 (353) | 1.00 (0.99-1.01)<sup>c</sup> | 0.69 | 1.00 (0.99-1.01)<sup>c</sup> | 0.70 |
| Wasted Work (mm Hg %) |  | 107 (50) | 117 (61) | 1.03 (0.96-1.11)<sup>c</sup> | 0.41 | 1.03 (0.95-1.11)<sup>c</sup> | 0.47 |
| Work Efficiency (%) |  | 94.6 (2.7) | 94.5 (2.4) | 0.99 (0.84-1.16) | 0.87 | 0.97 (0.81-1.15) | 0.69 |
| RV<br>Volume | Normal | 44 (96%) | 54 (98%) | Ref | 0.73 | Ref | 0.31 |
|  | Mildly Dilated | 1 (2%) | 1 (2%) | 2.45 (0.21-28.0)<sup>d</sup> |  | 3.55 (0.28-45.3)<sup>d</sup> |  |
|  | Moderately Dilated | 1 (2%) | 0 (0%) |  |  |  |  |
| RV<br>Function | Normal | 46 (100%) | 53 (96%) | Ref | 0.37 | Ref | 0.54 |
|  | Mildly Reduced | 0 (0%) | 2 (4%) | 0.23 (0.01-4.92)<sup>e</sup> |  | 0.38 (0.02-8.50)<sup>e</sup> |  |
| TAPSE (mm) |  | 22.5 (4.1) | 22.9 (3.6) | 1.03 (0.93-1.14) | 0.63 | 1.02 (0.91-1.14) | 0.75 |
| RV S' Velocity (cm/s) |  | 11.5 (2.1) | 11.6 (2.3) | 1.03 (0.85-1.24) | 0.76 | 1.05 (0.86-1.28) | 0.65 |
| RV Free Wall Longitudinal Strain<br>(%) |  | -23.6 (4.6) | -23.5 (4.6) | 1.01 (0.91-1.11) | 0.89 | 1.06 (0.94-1.18) | 0.37 |
| Cardiac Index (L/min/m<sup>2</sup>) |  | 2.5 (0.5) | 2.5 (0.5) | 1.18 (0.52-2.64) | 0.70 | 0.91 (0.35-2.39) | 0.85 |
| Pulmonary Artery Systolic<br>Pressure (mmHg) |  | 22.2 (4.1) | 22.9 (4.6) | 1.21 (0.69-2.12)<sup>f</sup> | 0.51 | 1.27 (0.64-2.55)<sup>f</sup> | 0.50 |
| Pericardial<br>Effusion | Trace/ Small | 4 (9%) | 0 (0%) | 12.0 (0.63-230)<sup>d</sup> | 0.10 | 8.11 (0.40-166)<sup>d</sup> | 0.11 |

Systolic Excursion; RV S'=right ventricular tricuspid annulus peak systolic velocity. Parameters are adjusted for age, sex, hospitalization, time since symptom onset, and medical history as described in the methods. <sup>a</sup>Per 5% decrease. <sup>b</sup>Per 5g/m<sup>2</sup> increase. <sup>c</sup>Per 10 mm Hg %. <sup>d</sup>Estimated using Firth Logistic Regression due to separation. <sup>e</sup>Mild & moderate combined due to small numbers. <sup>f</sup>Per 5 mm Hg increase.

**Figure 2.**
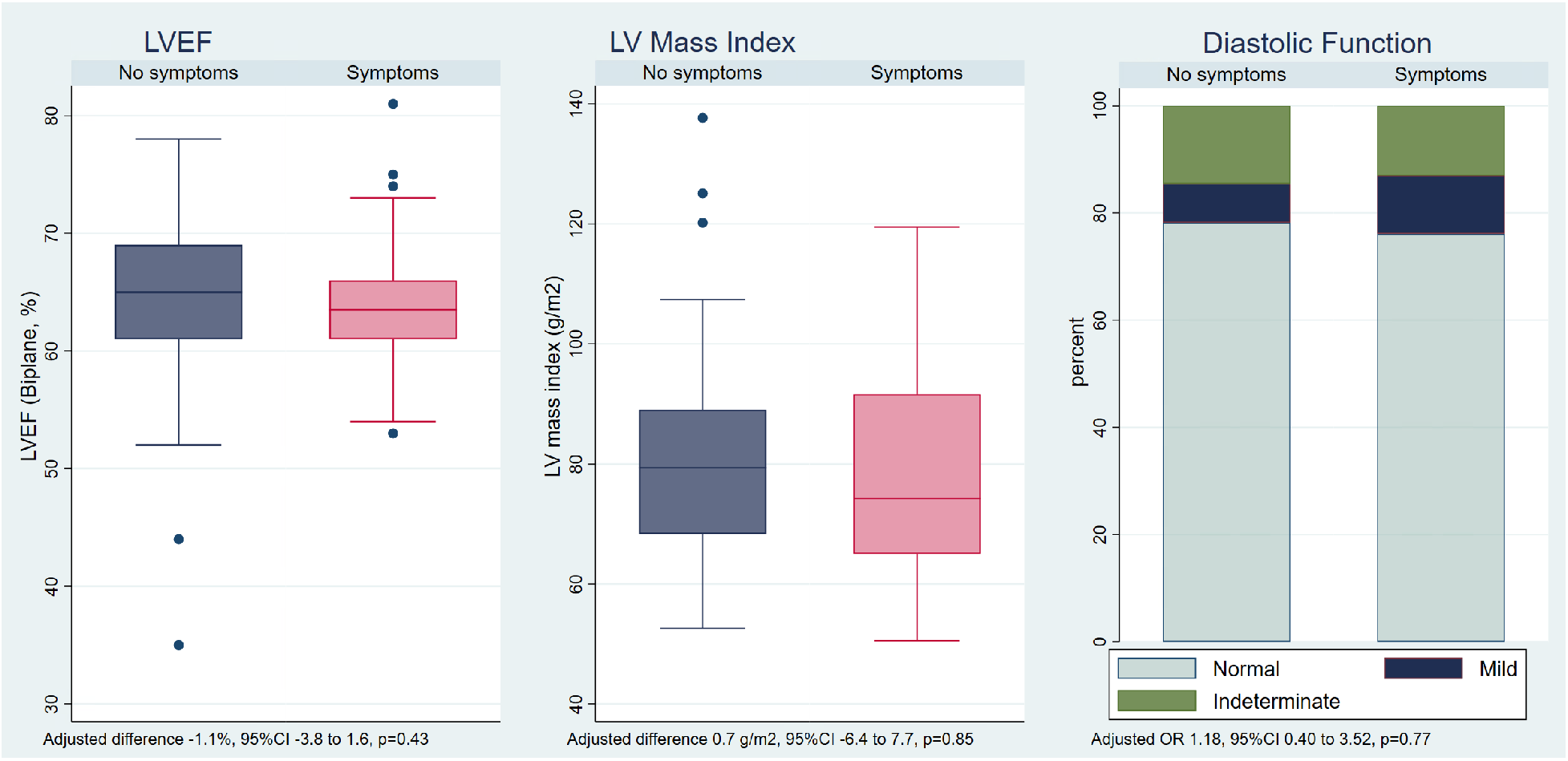
Left Ventricular Ejection Fraction, Mass Index, and Diastolic Function by Cardiopulmonary Symptoms. Boxplots of left ventricular ejection fraction (LVEF), left ventricular mass index, and bar plot of diastolic function. Odds of symptoms were 1.16 times higher per 5% decrease in LVEF, which was not statistically significant (95%CI 0.83-1.62; p=0.40). The odds of symptoms were not significantly higher with increased LV mass (1.01 per 5 g/m^2^, 95%CI 0.89-1.16; p= 0.81). 7% of those without symptoms and 11% with symptoms had mild diastolic dysfunction; the odds of symptoms were 1.77 times higher among those with diastolic dysfunction compared to those with normal diastolic function, which was not statistically significant but could not exclude a meaningful effect (95% CI 0.35-8.88; p=0.78).

**Table 3.** Associations between Echocardiographic Findings and Symptoms.

|  | Dyspnea,<br>Chest Pain,<br>or<br>Palpitations<br>(n=47) | Dyspnea<br>(n=33) | Chest Pain<br>(n=15) | Palpitations<br>(n=27) | Any<br>Potentially<br>Cardio-<br>pulmonary<br>Symptom<br>(n=64) | 2 or more<br>symptoms<br>(n=38) vs no<br>symptoms<br>(n=38) |
| --- | --- | --- | --- | --- | --- | --- |
| **LVEF, per**<br>**5% decrease** | 1.16<br>(0.83-1.62;<br>p=0.40) | 1.07<br>(0.76-1.52;<br>p=0.76) | 1.08<br>(0.67-1.72;<br>p=0.70) | 1.33<br>(0.87-2.03;<br>p=0.19) | 1.10<br>(0.78-1.53;<br>p=0.59) | 1.29<br>(0.87-1.89;<br>p=0.20) |
| **Presence of**<br>**Diastolic**<br>**Dysfunction** | 1.77<br>(0.35-8.88;<br>p=0.78) | 1.45<br>(0.29-7.36;<br>p=0.91) | 2.17<br>(0.45-7.30;<br>p=0.73) | 2.41<br>(0.34-17.3;<br>p=0.56) | 1.72<br>(0.34-8.76;<br>p=0.78) | 1.32<br>(0.20-8.73;<br>p=0.66) |
| **LV strain,**<br>**per 1% more**<br>**abnormal** | 1.07<br>(0.89-1.28; p<br>= 0.45) | 0.96<br>(0.80-1.16;<br>p = 0.67) | 1.23<br>(0.97-1.57;<br>p=0.09) | 0.96<br>(0.76-1.20;<br>p=0.70) | 1.01<br>(0.85-1.21;<br>p=0.90) | 1.03<br>(0.84-1.27;<br>p=0.75) |
| **Presence of**<br>**RV Dilation** | 3.55<br>(0.28-45.3;<br>p=0.33) | 1.27<br>(0.09-18.2;<br>p=0.86) | 3.35<br>(0.24-46.1;<br>p=0.37) | 2.29<br>(0.15-35.0;<br>p=0.55) | 1.98<br>(0.16-24.6;<br>p=0.60) | 1.72<br>(0.06-53.0;<br>p=0.76)* |
| **RV Strain**<br>**per 1% more**<br>**abnormal** | 1.05<br>(0.94-1.18;<br>p=0.38) | 0.99<br>(0.88-1.12;<br>p=0.89) | 1.12<br>(0.96-1.31;<br>p=0.14) | 0.91<br>(0.79-1.06;<br>p=0.22) | 1.07<br>(0.96-1.20;<br>p=0.22) | 1.07<br>(0.92-1.24;<br>p=0.40) |
| **Pulmonary**<br>**Artery**<br>**Pressure, per**<br>**5 mm Hg**<br>**increase** | 1.27<br>(0.64-2.55;<br>p=0.50) | 1.08<br>(0.51-2.28;<br>p=0.85) | 1.85<br>(0.78-4.37;<br>p=0.16) | 1.24<br>(0.55-2.81;<br>p=0.61) | 1.01<br>(0.50-2.03;<br>p=0.98) | 1.05<br>(0.42-2.61;<br>p=0.91) |
| **Presence of**<br>**Pericardial**<br>**Effusion\*** | 12.0<br>(0.63-230,<br>p=0.098) | 20.6<br>(1.07-394;<br>p=0.045) | 6.19<br>(0.98-39.1;<br>p=0.053) | 30.8<br>(1.59-594;<br>p=0.023) | 7.32<br>(0.38-140;<br>p=0.19) | 10.7<br>(0.55-206;<br>p=0.12) |

Other individual functional parameters including LV strain (OR 1.07, 95%CI 0.89-1.28; p = 0.45), RV strain (OR 1.05, 95%CI 0.94-1.18; p=0.38) and myocardial work (OR 1.00 per 10 mm Hg %, 95%CI 0.99-1.01; p=0.93) were not obviously associated with the composite outcome (Supplemental Figure 2). Hemodynamic markers including estimated cardiac index (OR 0.91 per L/min/m^2^, 95%CI 0.35-2.39; p=0.85) and pulmonary artery pressures (OR 1.27 per 5 mm Hg, 95%CI 0.64-2.55; p=0.50) were also not estimated to be substantially associated with symptoms, although a meaningful effect could not be ruled out. No participants had pulmonary hypertension (defined as a pulmonary artery systolic pressure over 35 mm Hg).

### Cardiac Biomarkers, Systemic Inflammatory Markers, and SARS-CoV-2 antibody levels

Individuals with cardiopulmonary symptoms had elevated hs-CRP compared to those without symptoms (median 1.8 mg/L with symptoms vs 0.9 mg/L without symptoms, p=0.03; Figure 3a and Table 4). For each doubling of hsCRP, the odds of having the composite outcome were 1.32 times higher (95%CI 1.01-1.73; p=0.02) and the odds of having two or more symptoms relative to no symptoms were 1.70 times higher (1.11-2.61; p=0.02). There was no estimated substantial association between hs-troponin and symptoms (OR 1.02 per doubling, 0.78-1.34; p=0.86), with the range of the confidence interval excluding a large clinical effect. The association between NT pro BNP and symptoms was estimated to be positive but inconclusive (OR 1.27, 95%CI 0.85-1.89; p=0.25).

**Figure 3.**
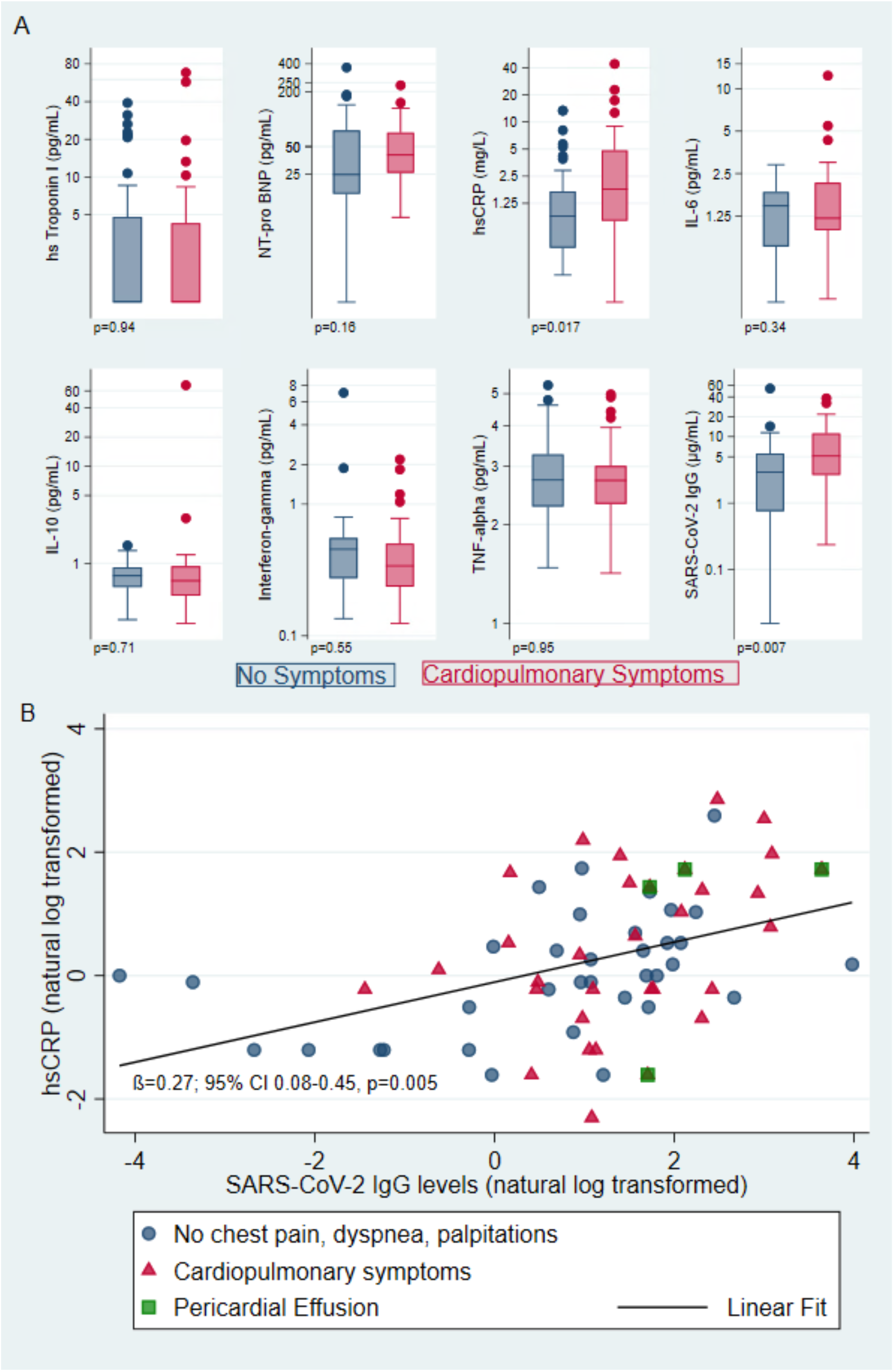
Biomarkers by Presence of Cardiopulmonary Symptoms and Relationship between Antibodies and hSCRP. Panel A shows box and whisker plots of biomarkers plotted on log-scale including hs-troponin I, NT-pro-BNP, hs-CRP, IL-6, IL-10, IFNγ, TNFα, and SARS-CoV-2 receptor binding domain IgG antibodies, with p-values listed for unadjusted t-tests of log-transformed markers. For each doubling of hsCRP, the odds of having dyspnea, chest pain or palpitations were 1.32 times higher (95%CI 1.01-1.73; p=0.02) and 1.42 times higher per doubling of antibody levels (95% CI 1.06-1.90; p=0.02). Other biomarkers were not strongly associated with symptoms. Panel B demonstrates that natural log transformed antibody levels and hsCRP are correlated (adjusted ß=0.27; 95% CI 0.08-0.45 p=0.005). Both antibody levels and hsCRP are higher in those with symptoms (red triangles) than those without symptoms (blue circles). The association between antibody levels and hsCRP did not vary by symptom status (p for interaction=0.51 and minimal change in beta coefficient). Those with pericardial effusions (green squares) had higher antibody levels and higher hsCRP.

**Table 4.**
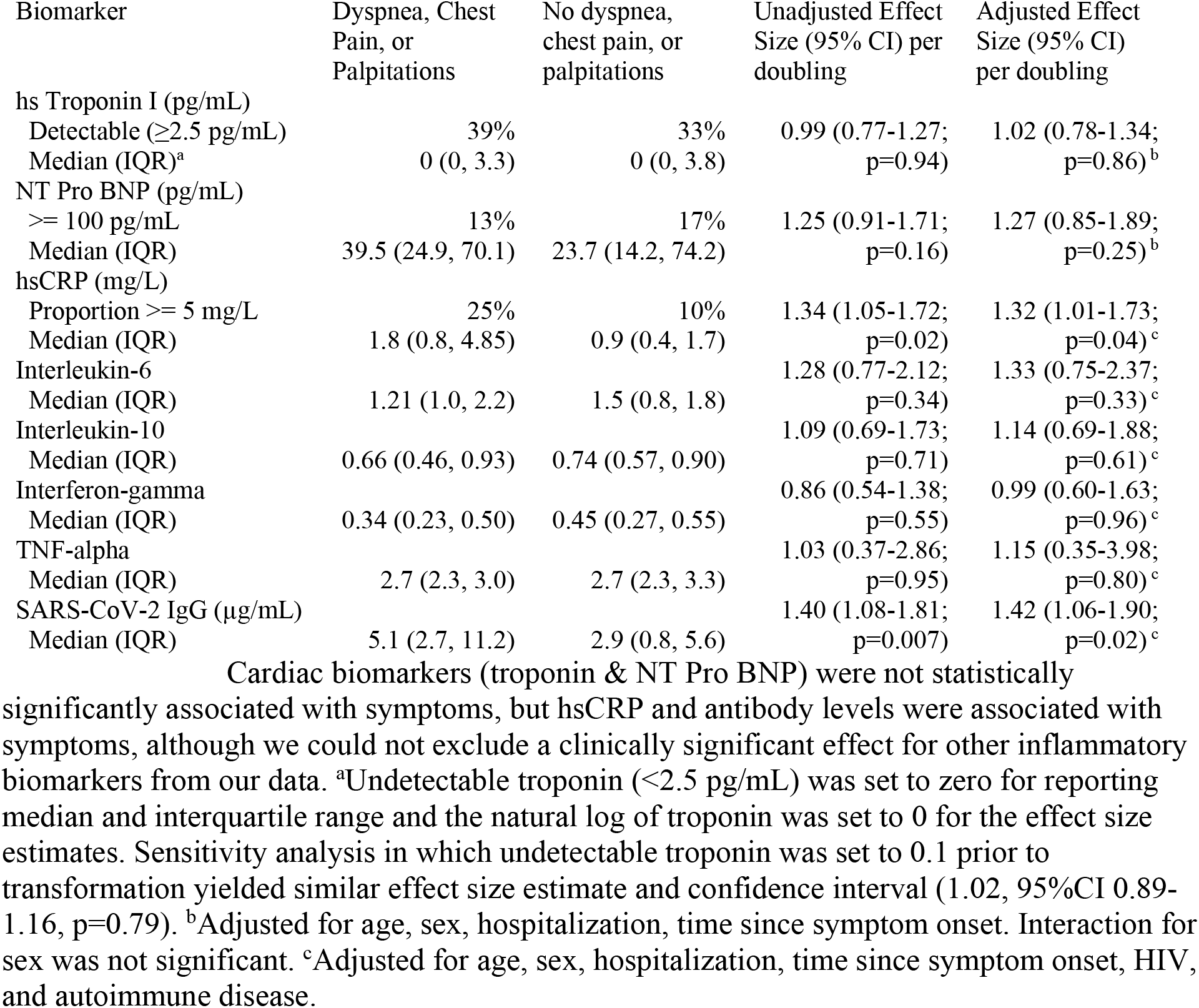
Associations between Biomarkers and Symptoms.

Additional inflammatory biomarker and antibody data were available for 73/102 (72%) participants. In this subset, we observed higher SARS-CoV-2 receptor binding domain IgG antibody levels among those with symptoms compared to those without symptoms (median 5.1 vs 2.9 µg/ml; p=0.02). The odds of having the composite outcome were 1.42 times higher per doubling of antibody levels (95%CI 1.06-1.90; p=0.02) and the odds of having 2 or more symptoms were 1.39 times higher (95%CI 0.96-2.01; p=0.09). Although the odds of having the composite outcome were not statistically significantly higher per doubling of IL-6 (OR 1.33, 95%CI 0.75-2.37; p=0.33), the odds of having 2 or more symptoms versus no symptoms were 4.01 times higher per doubling of IL-6 (95%CI 1.20-13.2; p=0.02). Results for IL-10, IFNγ, and TNFα did not demonstrate any significant associations with the composite outcome (Table 4) and having 2 or more symptoms (data not shown).

The odds of having a pericardial effusion were estimated to be 1.98 times higher per doubling of IL-6 (95%CI 0.85-4.63; p=0.12), 1.35 times higher per doubling of hsCRP (95%CI 0.75-2.43; p=0.32), and 1.87 times higher per doubling of SARS-CoV-2 antibody levels (95%CI 0.92-3.81; p=0.09) though none of these results reached statistical significance. No individuals with pericardial effusions had a high sensitivity troponin >5 pg/ml at the time of the echocardiogram suggestive of ongoing myocarditis. Biomarkers among those with and without pericardial effusions are shown in Supplemental Figure 3.

### Correlation between antibody levels and hsCRP

We found a statistically significant linear correlation between log-transformed antibody levels and hsCRP (adjusted ß=0.27; 95% CI 0.08-0.45; p=0.005; Figure 3b). The association between antibody levels and hsCRP did not vary by symptom status (p for interaction=0.51 and minimal change in beta coefficient). Antibody levels are correlated with IL-6 levels only among those with symptoms (ß=0.25; 95% CI 0.001-0.51; p=0.05; Supplemental Figure 4).

## Discussion

In this cross-sectional analysis of a prospective COVID-19 recovery cohort, we found that higher antibody levels, markers of inflammation (hsCRP and possibly IL-6), and possibly pericardial effusions were associated with cardiopulmonary PASC at a median 7 months after infection with SARS-CoV-2. Taken together, these data suggest that symptoms in cardiopulmonary PASC are not due to myocardial injury or changes in cardiac function, but rather associated with higher antibody levels and a systemic inflammatory process. To our knowledge this is the first study to demonstrate the association between the presence of pericardial effusions and inflammatory biomarkers in the setting of PASC. Our findings are an important first step in understanding the mechanism of PASC which will be critical to treat and prevent this complication which affects a meaningful proportion of individuals following SARS-CoV-2 infection.

### Determinants for Cardiopulmonary PASC

The two patient characteristics strongly associated with ongoing cardiopulmonary symptoms were female sex and hospitalization for acute COVID-19. Female sex has been previously associated with COVID-19 symptoms lasting greater than 28 days (22), 6 months (23), and one year (3). Several studies have previously demonstrated that severity of initial illness, and specifically hospitalization, are associated with symptoms at 30 and 60 days and at 6 months (23, 24).

### Antibody levels and Inflammation in Cardiopulmonary PASC

We demonstrated an association between higher antibody levels and elevated markers of inflammation, namely hsCRP, with cardiopulmonary symptoms, which supports the role of inflammation as a putative mechanism underlying cardiopulmonary PASC rather than myocardial injury resulting in abnormal cardiac function. Our findings are consistent with prior studies that found higher antibody levels among those with symptoms at 6 months (23, 25), although a separate study found lower antibody levels but higher CRP among those with the most symptoms (26). Our study is also consistent with a prior study that found normal high sensitivity troponin and NT-proBNP levels among individuals 6 months after mild infection compared to matched uninfected controls (8).

Higher levels of antibodies among those with symptoms could reflect more severe initial infection (25) or possibly viral antigen persistence (27), both of which have been suggested as contributors to PASC; either process could be related to persistent inflammation we found. The identification of pericardial effusions among a subset with symptoms and elevated antibody levels, hs-CRP, and IL-6 among those individuals raises the possibility that localized organ inflammation may be present beyond the early convalescent phase (2-3 months) where pericardial effusions have been noted in CMR studies (5, 6, 28). Troponin was low or undetectable among all those with pericardial effusions, providing strong evidence against ongoing myocardial injury or active myopericarditis. Whether or not this phenomenon is distinct from post-viral pericarditis (29), well-described after other viruses, is uncertain. The presence of pericardial effusions following COVID-19 is similar to pericardial effusion in the setting of HIV before the antiretroviral era (30).

### Lack of evidence supporting cardiac structural or functional pathology underlying PASC

Half of those with cardiopulmonary symptoms attributed to COVID had no structural or functional abnormalities evident on echocardiography, and the functional abnormalities present among the remainder did not provide important clues to the etiology of symptoms with the possible exceptions of diastolic function and RV dilation. Our findings provide strong evidence that active ongoing myocarditis leading to heart failure or persistent pulmonary hypertension from residual lung injury are not common pathways to symptoms among the majority of those with cardiopulmonary PASC. Other organic cardiac pathologic changes that may underlie cardiopulmonary PASC that are not detectable by resting echocardiography include ischemia including due to microvascular dysfunction (6), abnormal diastolic function or pulmonary hypertension with exertion (31), and autonomic dysfunction due to nerve involvement (11, 32).

Our study is consistent with previously published echocardiographic studies in early COVID-19 convalescence. For example, several studies found a high prevalence of persistent cardiopulmonary symptoms lasting up to 1-3 months after hospitalization for acute infection, but a low prevalence of echocardiographic abnormalities and normal LV and RV function (33-35). Two studies performed TTE 6 months after hospitalization for COVID-19 in a total of 94 individuals and found no rest abnormalities and no differences between those with and without myocardial injury during acute infection (31, 36). Our study extends upon these findings by including individuals not hospitalized for severe COVID-19 and demonstrating that significant cardiac functional changes are not common >6 months following acute SARS-CoV-2 infection, and including assessment of antibody titers and inflammatory markers.

Similarly, our findings are consistent with cardiac MRI structural and functional findings with normal LV and RV function the predominant findings (4, 6, 28). Although several studies found evidence of inflammation and late gadolinium enhancement suggestive of fibrosis early after acute SARS-CoV-2 recovery in a high proportion of study participants, the long-term implications of these findings remain uncertain and will require further investigation. Other studies have suggested that myocarditis and cardiac fibrosis after COVID-19 may be rare, with a prevalence less than 1% among young healthy athletes (37, 38) and no difference between healthcare workers with mild SARS-CoV-2 infection compared to controls (8).

### Therapeutic Implications: Cardiac evaluation & Anti-inflammatory Therapy for PASC

Given the millions of individuals who have been infected with COVID-19 globally, and emerging reports of persistent symptoms in a meaningful proportion of individuals (39-43), studies to identify and ultimately treat or prevent PASC are of the utmost importance. Clinical evaluation and management for individuals with suspected cardiopulmonary PASC is unknown but could include biomarker measurement, electrocardiogram and echocardiogram and should be tailored to each patient’s clinical scenario, particularly to rule out non-COVID pathology and to target consideration for advanced testing such as CMR or cardiopulmonary exercise testing. With regards to therapeutic strategies, different anti-inflammatory approaches have had mixed results in the setting of hospitalized COVID-19 (44-51) as well as outpatients with COVID-19 (52). Mechanistic studies with long-term follow-up are needed to elucidate the role of inflammation in PASC. Our findings suggest that anti-inflammatory therapies and/or monoclonal antibodies may be reasonable strategies to alleviate or prevent persistent cardiopulmonary symptoms following COVID-19, which will need to be carefully evaluated in future clinical trials.

### Study Limitations

Limitations of this study include the use of a convenience sample and the cross-sectional echocardiographic and biomarker assessments. There is a risk of selection bias from those in the LIINC study who chose to participate in the cardiovascular sub-study, and from the shift in our recruitment criteria toward those with symptoms. To date, there are no formal definitions of cardiopulmonary PASC. We did not have echocardiograms from prior to or during acute infection to examine sub-clinical changes among our sample. Because we are specifically interested in the pathophysiology of persistent symptoms compared to those who fully recover from COVID-19, we intentionally did not include a SARS-CoV-2 uninfected control group, but inclusion of such a control group would have strengthened our inferences. We excluded those with pre-existing heart failure, congenital heart disease, and pulmonary hypertension, so our findings may not be generalizable to those with pre-existing cardiac disease. Third, because we only included a small number of people who received intensive care during acute COVID-19 and none with myocarditis in the setting of acute disease, our findings may not be applicable to those with the highest severity of illness. Finally, the number with pericardial effusions is small so findings particularly with respect to biomarkers and pericardial effusions should be confirmed in larger studies.

## Conclusions

In conclusion, SARS-CoV-2 antibody levels, inflammatory biomarkers, and possibly pericardial effusions were associated with cardiopulmonary symptoms, but other echocardiographic structural and functional parameters were not associated with PASC (with possible exceptions of diastolic dysfunction and RV dilation, which were inconclusive). Further studies into mechanisms of elevated antibody levels and increased inflammation in PASC will provide much needed insight into therapeutic targets in this disease process.

## Data Availability

All data produced in the present study are available upon reasonable request to the authors.

## Abbreviations list

TTE: transthoracic echocardiogram
PASC: (post-acute sequelae of COVID-19)
hsCRP: high sensitivity c reactive protein
LIINC: Long-term Impact of Infection with Novel Coronavirus
HIV: human immunodeficiency
NT-pro-BNP: N-terminal prohormone b-type natriuretic protein
IL: interleukin
TAPSE: tricuspid annular plane systolic excursion
POTS: postural orthostatic tachycardia syndrome
LV: left ventricle
RV: right ventricle

## Author Contributions

MSD designed the study, acquired and analyzed the data, and wrote the first draft of the manuscript. MJP designed the study, acquired data, provided critical input on data interpretation, and provided critical input on the manuscript. JDK helped with data interpretation and critical edits to the manuscript. SW oversaw echocardiographic measurements and provided critical input to the manuscript. SS, DL, VMA, VZ, KS, CH, MIA, RH, VT acquired data and provided feedback on initial drafts of the manuscript. SL participated in data management and analysis. SShao participated in acquiring data and designed the graphical abstract. AC, BCY, JWW and CJP measured biomarkers and antibodies and provided input on the manuscript. JK helped design the statistical plan, data analysis and interpretation, and critical edits to the manuscript. JDK, TJH, JNM, SGD, and PYH provided critical input on study design, study conduct, data interpretation, and edits to the manuscript.

## Acknowledgements

We are grateful to the study participants. We are grateful to the clinical staff caring for these individuals. We would like to thank Dr. Kara Lynch and Dr. Alan Wu for their assistance with measuring hs-troponin, NT-Pro-BNP and hs-CRP. We would also like to acknowledge support from Jeremy Lambert from Quanterix. We acknowledge the contributions of the UCSF Clinical and Translational Science Unit.

---

**Supplemental Table.**
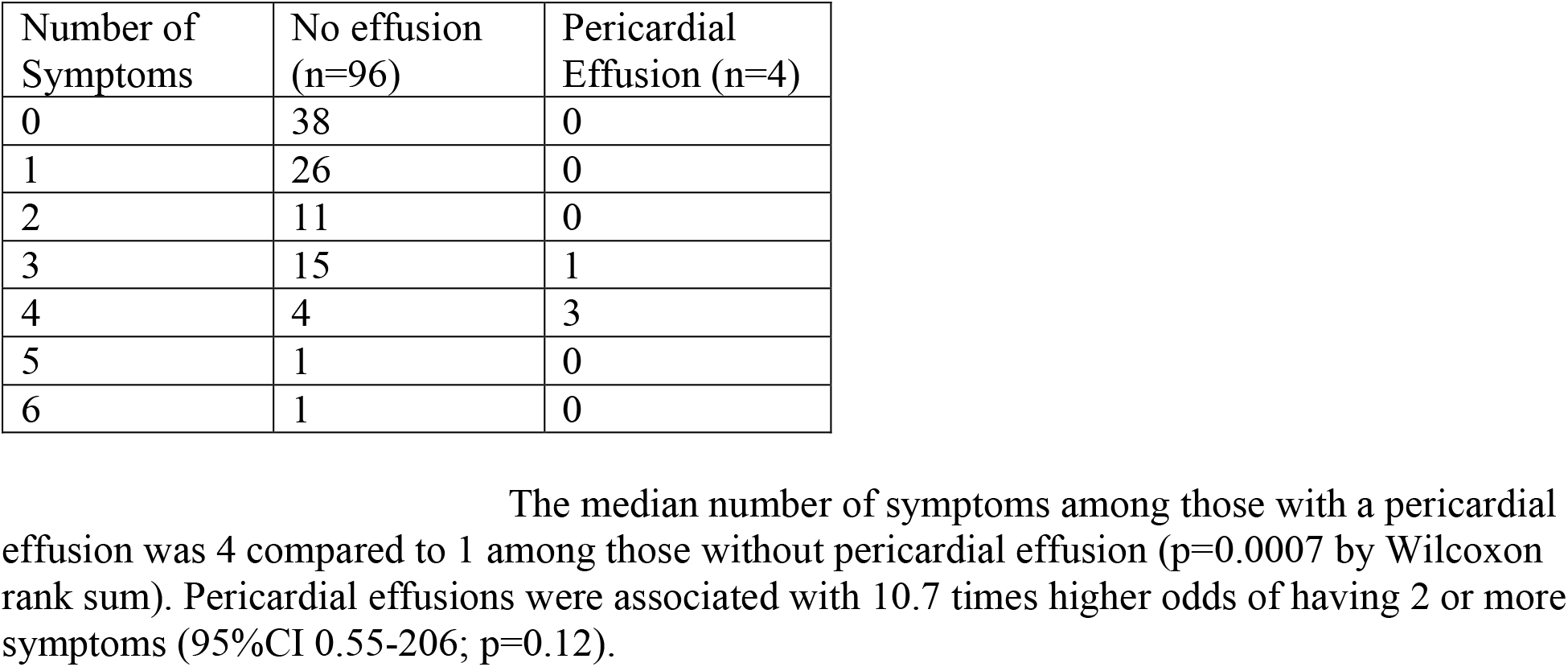
Number of potentially cardiopulmonary symptoms among those with and without pericardial effusion.

**Supplemental Figure 1.**
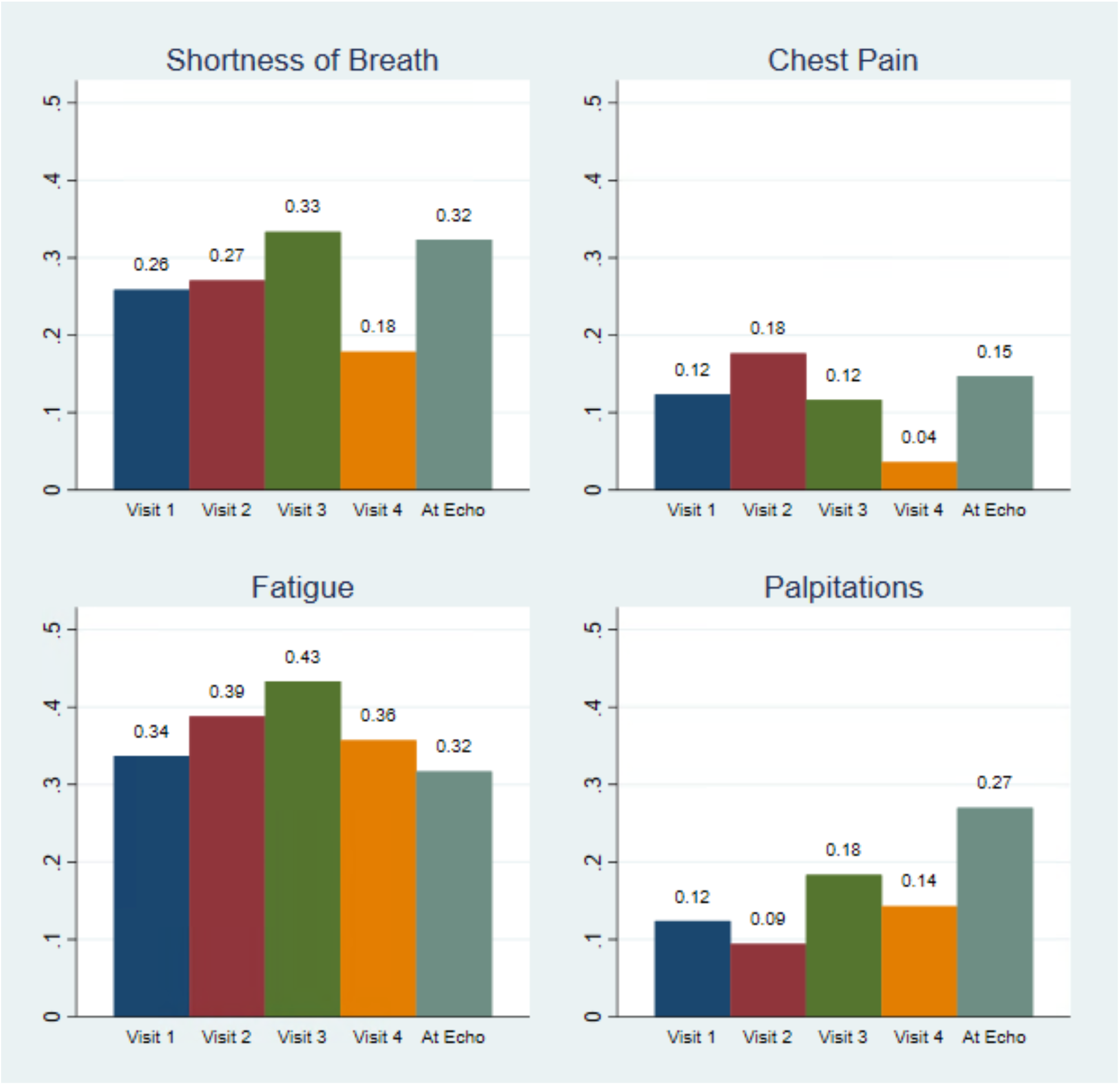
Persistence of cardiopulmonary symptoms over time. The four panels show the prevalence of shortness of breath, chest pain, fatigue, and palpitations at each study visit; note that these are specific to the study sample and do not represent the true prevalence of these symptoms among all those recovering from COVID-19, but rather demonstrate that the single time point that we present the symptoms is a reasonable approximation to assess the association between biomarkers and echocardiograms. Note that we still adjusted our analyses for time since COVID-19 symptom onset or positive PCR test with the exception of baseline characteristics present at onset of acute infection. Visit 1 took place at a median of 2 months (N=89), Visit 2 at 3 months (N=85), Visit 3 at 4.5 months (N=60), and Visit 4 at 9 months (N=28).

**Supplemental Figure 2.**
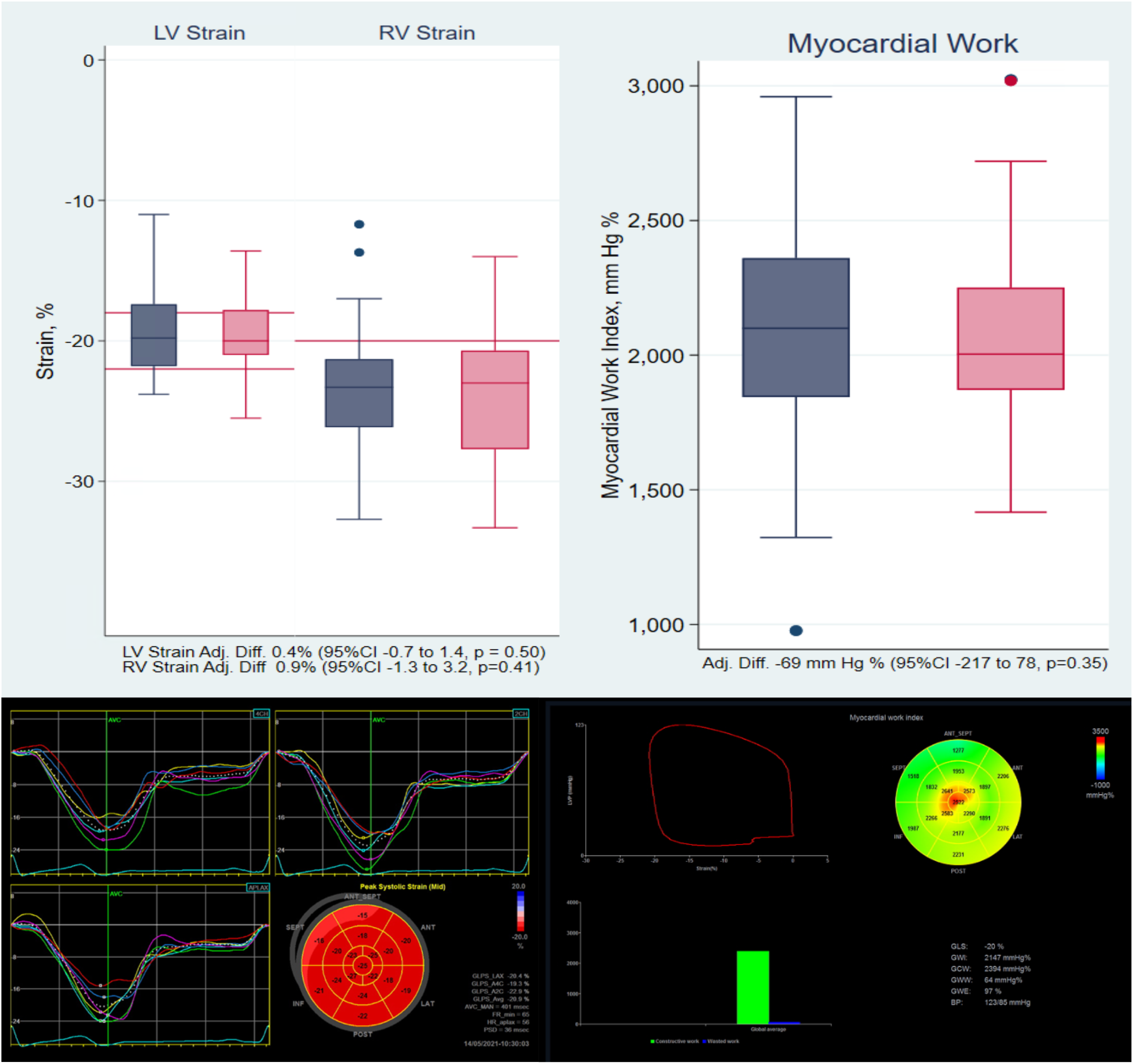
Strain and Myocardial Work by Cardiopulmonary Symptoms. Boxplots of left ventricular peak global longitudinal strain (normal -18 to -22%, red lines),right ventricular strain (normal < -20%, red line), and myocardial work among those without symptoms (navy blue) and with symptoms (pink) are shown on the top. LV strain (OR 1.07, 95%CI 0.89-1.28; p = 0.45), RV strain (OR 1.05, 95%CI 0.94-1.18; p=0.38), and myocardial work (OR 1.00 per 10 mm Hg %, 95%CI 0.99-1.01; p=0.93) were not associated with symptoms. Representative left ventricular strain tracings and myocardial work strain pressure curves and regional myocardial work are shown below.

**Supplemental Figure 3.**
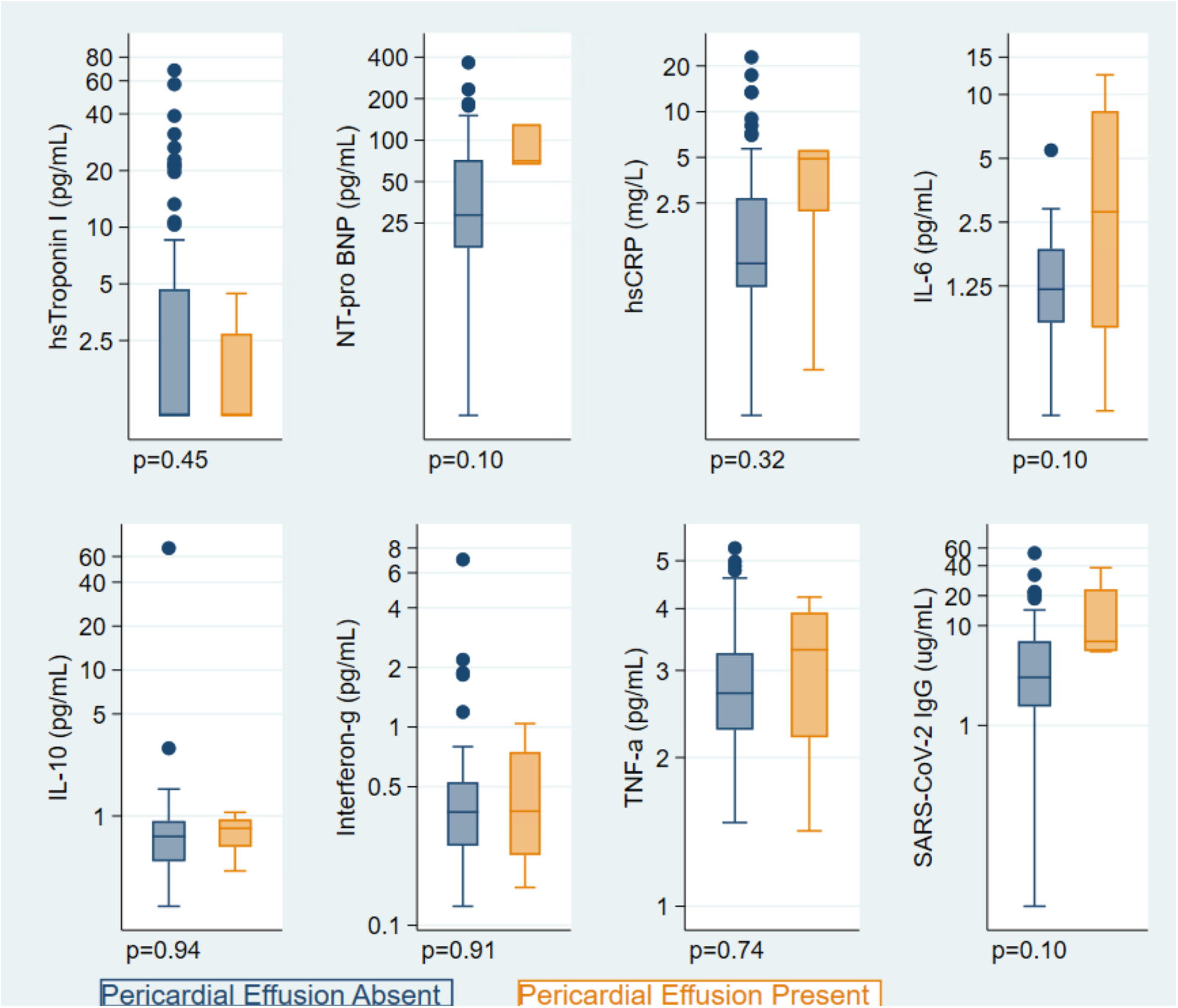
Biomarkers by Presence of Pericardial Effusion. Box and whisker plots of biomarkers by presence of pericardial effusion plotted on log-scale including hs-troponin I, NT-pro-BNP, hs-CRP, IL-6, IL-10, Interferon-gamma, TNF-alpha, and SARS-CoV-2 IgG antibodies. With only 4 individuals with pericardial effusions, the statistical power is low, but our findings suggest higher levels of NT-pro BNP, hsCRP, IL-6, and antibodies may be present among those with pericardial effusions.

**Supplemental Figure 4.**
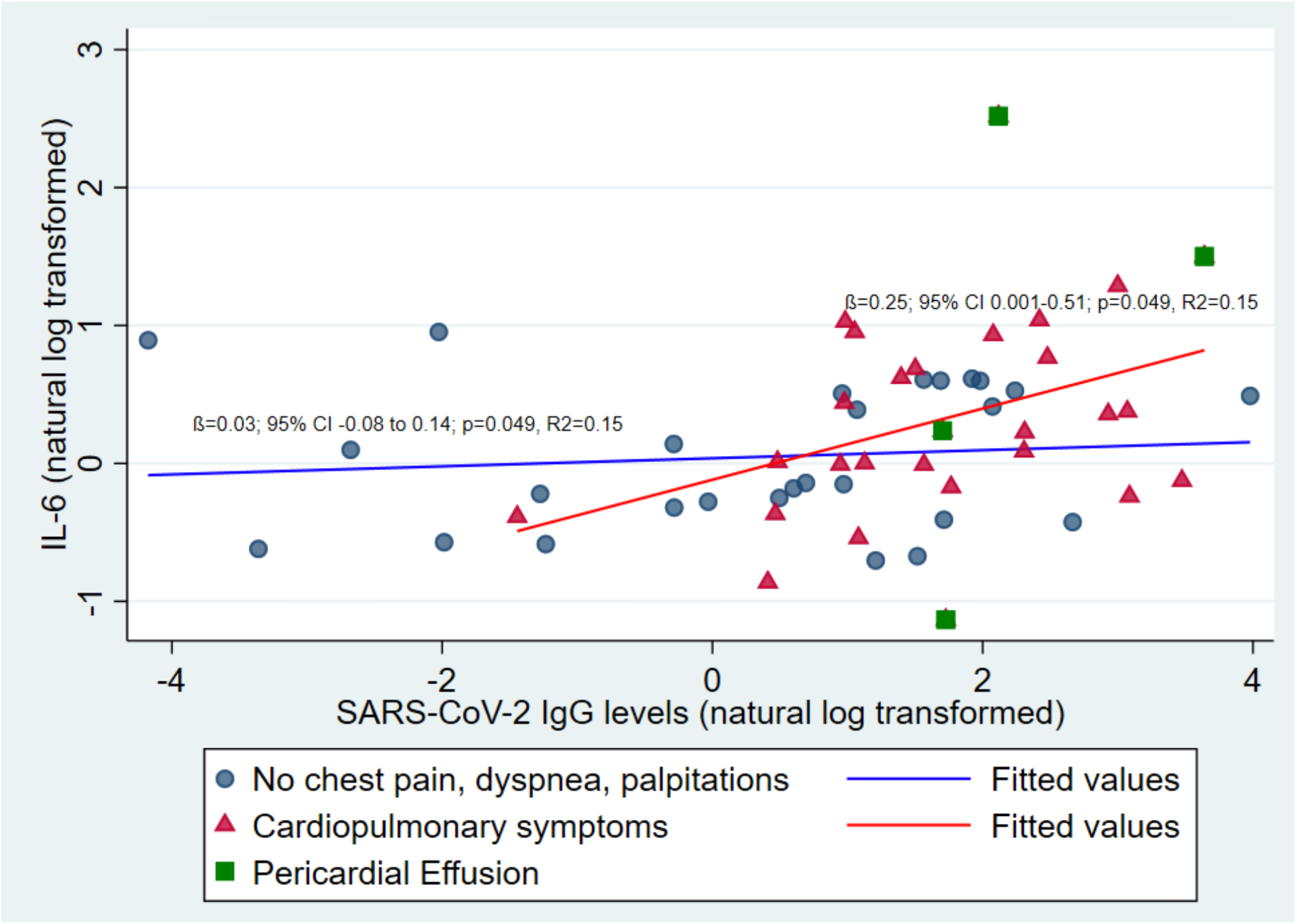
Relationship between Antibody levels, IL-6, Symptoms and Pericardial Effusions. Antibody levels are correlated with IL-6 levels only among those with symptoms (red triangles & trend line; ß=0.25; 95% CI 0.001-0.51; p=0.049, R^2^=0.15), but not among those without symptoms (blue circles & trend line; ß=0.03; 95% CI -0.08 to 0.14; p=0.049, R^2^=0.15). Individuals with pericardial effusions are shown as green squares.

